# ClinSV: Clinical grade structural and copy number variant detection from whole genome sequencing data

**DOI:** 10.1101/2020.06.30.20143453

**Authors:** Andre E Minoche, Ben Lundie, Greg B Peters, Thomas Ohnesorg, Mark Pinese, David M Thomas, Andreas Zankl, Tony Roscioli, Nicole Schonrock, Sarah Kummerfeld, Leslie Burnett, Marcel E Dinger, Mark J Cowley

## Abstract

Whole genome sequencing (WGS) has the potential to outperform clinical microarrays for the detection of structural variants (SV) including copy number variants (CNVs), but has been challenged by high false positive rates. Here we present *ClinSV*, a WGS based SV integration, annotation, prioritisation and visualisation method, which identified 99.8% of pathogenic ClinVar CNVs >10kb and 11/11 pathogenic variants from matched microarrays. The false positive rate was low (1.5–4.5%) and reproducibility high (95–99%). In clinical practice, *ClinSV* identified reportable variants in 22 of 485 patients (4.7%) of which 35–63% were not detectable by current clinical microarray designs.

## Background

Genomic structural variant(s) (SV(s)), including copy number variant(s) (CNV(s)), are an important source of genetic variation, and it is well established that large CNVs (typically >100 kb) are an important cause of many inherited human genetic diseases [1–3]. Clinically accredited array comparative genome hybridization (aCGH), or single nucleotide polymorphism (SNP) microarrays (MA) are currently first-line clinical laboratory tests used to diagnose patients with many rare genetic diseases. Especially common among these conditions are intellectual disability [4] and autism [5]. Depending on their probe density, aCGH and SNP MA detect CNV down to a resolution of ∼50 kb and are unable to detect copy number-neutral SV events, such as inversions or balanced translocations. Short-read whole genome sequencing (WGS) has emerged as a comprehensive test for diagnosing rare inherited genetic disorders. A small number of laboratories have obtained clinical accreditation (e.g. CLIA/CAP [6], or ISO 15189 [7]) for the identification of single nucleotide variants (SNVs) and short insertion or deletion variants (indels) (<50bp) from WGS, where sensitivity and specificity often exceed 99% across most of the genome. Due to its broad and uniform sequencing coverage, WGS additionally has considerable potential to identify both small and large CNVs, with size ranging from smaller-than-single-exon, up to whole-chromosome aneuploidy, as well as copy-number-neutral SVs. Until recently, WGS analytical methods had imperfect sensitivity and specificity for detection of SVs, leading some groups to report that SV detection from WGS was not yet fit for use in clinical practice [8,9]. There are now numerous recent reports using WGS to identify short CNV, affecting single genes, or even single exons as the cause of many monogenic disorders, suggesting considerable potential for short CNVs below the limit of detection of traditional MA to explain a proportion of previously undiagnosed patients [4,10,11]. The ability to find short, potentially pathogenic CNVs raises significant new interpretation challenges, as there are thousands of benign short CNVs in healthy individuals [12,13].

For the detection of SVs and CNVs, short-read paired-end sequencing provides three main categories of evidence: changes in depth of coverage (DOC), reads with a gapped sequence alignments referred to as split reads (SR) and reads-pairs with aberrant mapping orientation or distance referred to as discordant pairs (DP, Fig. 1a, 1b). Most CNV detection tools (e.g., CNVnator [14]) use only changes in DOC, while most popular SV detection tools (e.g., Lumpy [15], Delly2 [16], Manta [17]) use evidence from SP and DP to identify SVs [16,18–20]. Used alone, each of these methods gives an incomplete picture. DOC methods work well for large CNVs with confidently mapped reads but perform poorly for shorter CNVs, and for regions with poorly mapped reads, or extreme GC content. Recent improvements in DOC methods have demonstrated high quality CNVs calling from WGS down to 10 kb [20]. SR and DP can identify small CNVs and copy number-neutral SVs, often with base-pair precision, but perform poorly when supporting reads are ambiguously aligned, and suffer from reduced coverage at regions of high GC nucleotide composition. Previous studies have proposed to improve CNV and SV detection by integrating multiple variant callers [21–24]. These approaches often combine large numbers of individual CNV and SV callers to increase sensitivity, at the expense of specificity, complexity and analytical cost, and do not address the challenges of variant visualization, annotation, and prioritization of rare variants.

**Figure 1.**
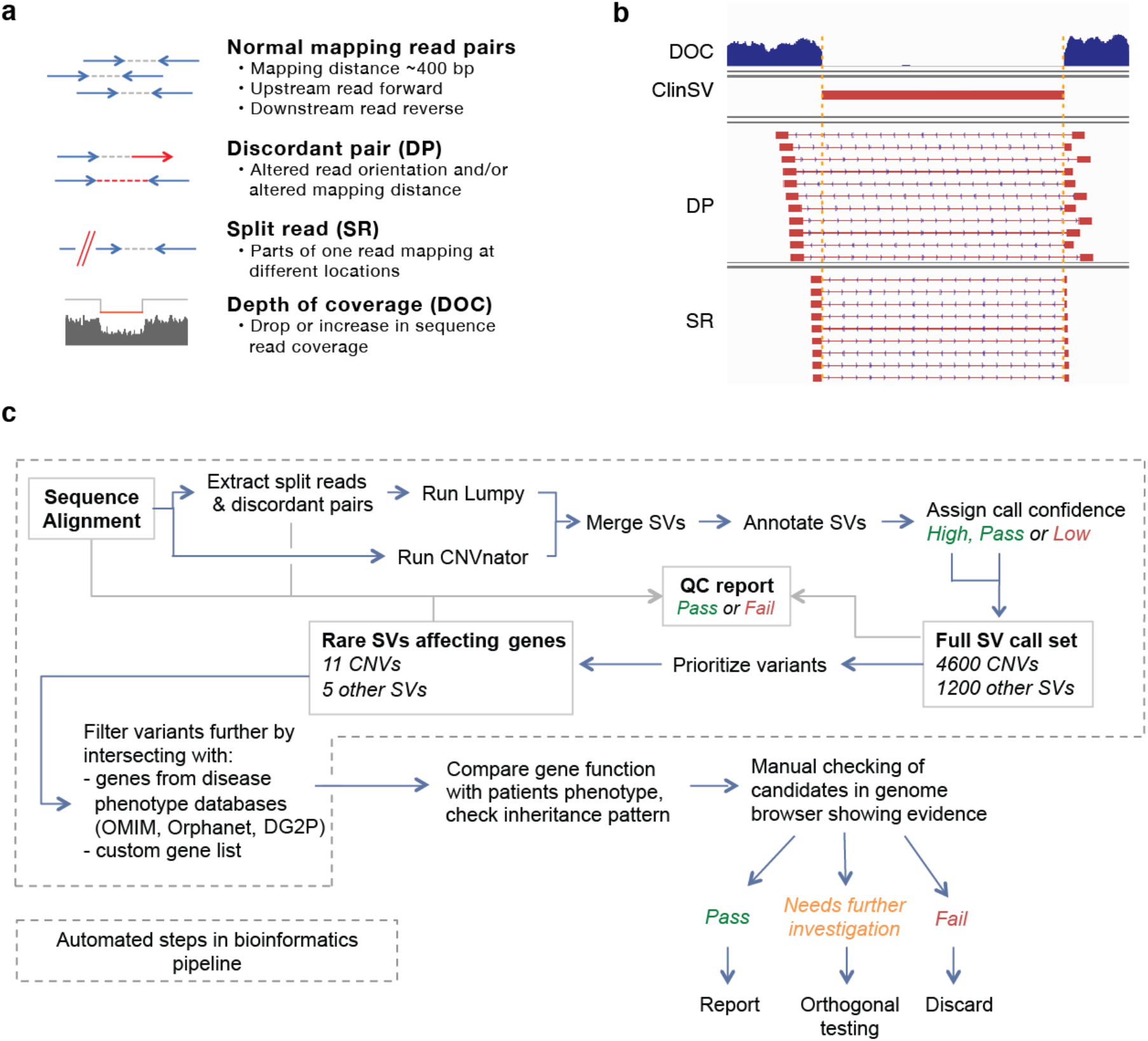
Evidence types used for CNV and SV identification, and the *ClinSV* workflow. **A:** The different evidence types used for automated SV identification from short-read paired-end WGS data. **B:** An example homozygous CNV deletion with clear breakpoints demarcated by a sharp drop in read coverage, and high numbers of supporting split reads (SR) and discordant read pairs (DP). **C:** The *ClinSV* workflow demonstrates the integration of two variant callers, variant annotation, quality classification, prioritization and interpretation.

Manual assessment of individual candidate SVs remains critical, with many groups using Integrated Genome Viewer (IGV) [25], or more specialized tools including svviz [26], SplitThreader [27], Ribbon [28], or SVPV [29]. However, most of these tools are limited to visualizing read evidence from single samples, ignoring additional lines of evidence to assess the quality, and clinical relevance of each candidate SV.

The variants that cause inherited genetic diseases are typically very rare in the general population. Deriving a database of SV frequencies however is challenging, as there is currently no consensus on the method or technology used to detect CNVs or SVs from genomic data. Furthermore, most databases of CNVs, such as the database of genomic variants (DGV) [30], include predominantly MA studies with varying resolutions making it difficult to determine whether partially overlapping CNVs represent the same event. Very recently, the most comprehensive resource of small and large CNVs derived from WGS is from the Genome Aggregation Database (gnomAD) [31], which was established using numerous CNV and SV callers.

Assessing the analytical performance of new SV detection methods is hampered by the lack of comprehensive and accurate reference call sets. While several validated SV datasets using the NA12878 reference cell line exist [15,26,32,33], the concordance between three of them was only 33% [34]. This poor concordance is likely due to differences in the technology (e.g. MA, WGS, PCR) and analytical approaches used. More recently, long-read sequencing datasets provide more comprehensive gold standard call sets, which can now be used for calibration and benchmarking [33,35].

To overcome the challenges associated with accurate CNV and SV detection for rare disease diagnosis, we have developed *ClinSV*, a computer-assisted analysis and visualization framework. We developed and optimized *ClinSV* through extensive *in silico* benchmarking experiments and comparisons to patients with matched MA from clinical laboratories. Through the analysis of 500 healthy elderly controls with 30–40x WGS, we compiled a population allele frequency database of CNV and SV, to assist with prioritizing rare, pathogenic CNV. *ClinSV* received ISO15189 clinical accreditation from National Association of Testing Authorities, Australia (NATA) in March 2018 and here we report the use of *ClinSV* across 485 consecutive patients referred for WGS within a clinically accredited diagnostic genomics laboratory.

## Methods

### Reference materials, patient recruitment and ethical compliance

DNA was obtained from the GIAB consortium from the NA12878 (GIAB1) and Ashkenazi trio (GIAB2) reference cell lines. DNA was provided for 11 patients from cohort A and 17 patients from cohort B whom were submitted for diagnostic testing at two Australian clinical laboratories for clinical microarray analysis. These samples were provided solely for the purpose of methods development and evaluating CNV performance, and do not have consent for the release of raw or processed genomic data. 485 patients were referred to the Genome.One diagnostic laboratory for genomic testing (including CNV analysis) in a clinical setting, and do not have consent for the release of raw or processed genomic data. WGS data from 500 healthy controls were provided by the Medical Genomics Reference Bank (MGRB) [36,37]. This research was approved by the ethics committee from the Royal Prince Alfred Hospital, Sydney, Australia (HREC ref no. 13/RPAH/363).

### Genome sequencing and primary analysis

DNA samples were processed using standard Illumina TruSeq Nano HT v2.5 library preparation on Hamilton Star instruments and sequencing was performed on Illumina HiSeq X instruments at the Kinghorn Centre for Clinical Genomics at Garvan Institute of Medical Research, Sydney, Australia. Only samples passing sequencing QC were processed further, i.e. mean coverage > 30x (coverage calculation according to Illumina technical note), >75% Q30 bases and total yield >100 Gb. Sequence reads were aligned to the human genome reference assembly GRCh37 decoy using BWA MEM (v0.7.12-r1039, settings -M).

### ClinSV pre-processing analysis

Discordant mapping read pairs (DP) and split reads (SR) were extracted from the sequence alignment file. To determine the upper and lower discordant mapping distance cut-offs, the mapping distance for all read pairs with normal mapping orientation in a sample region (1:1,000,001-5,000,000) was obtained by *ClinSV*. The read pairs were sorted by their mapping distance. Iterating through this list in descending order and summing the number of read-pairs, the upper cut-off was set to the mapping distance at an average coverage of 100 read pairs per Mb. The lower limit was obtained accordingly by iterating in ascending order. DP were extracted from the whole BAM, using PE-SR-bam2bed.pl (part of *ClinSV*) and these cutoffs. SR were extracted using the ‘extractSplitReads_BwaMem’ script from Lumpy [15] and further processed with PE-SR-bam2bed.pl, discarding reads mapping to NC_007605 or hs37d5, reads having more than one clipped part, multiple alignments or overlapping alignment parts. Coverage was assessed using the get_coverages.py script, and an exclude regions file was generated to exclude regions with >300x coverage using the get_exclude_regions.py scripts from Lumpy. Lumpy v0.2.11 was run to detect genome-wide SV with the options -mw 3 -tt 0, using only the discordant and split reads. Resulting variant calls were annotated with depth of coverage (DOC, see section “Depth of coverage annotation” below). When joint-calling multiple samples, deletions and duplications lacking DOC support in one or multiple samples were outputted as separate balanced variant calls (CNV=0).

CNVnator v0.3 was run in parallel using cnvnator_wrapper.py v.0.0.1 [21] with a window size of 100 bases. CNVnator calls spanning masked regions (marked with Ns in FASTA) of the genome were split, excluding the masked part. CNVnator calls were annotated with DP and SR showing the expected mapping orientation [15] and being located within certain inner (towards the center of the CNV) and outer limits (flanking the CNV) of each breakpoint. The outer limit was set to 1% of CNV length but at least 1 kb and at most 5 kb. The inner limit was set to half the CNV length but at most 5 kb.

The genotype was assigned purely based on the DOC, since the percentage of DPs and SRs were found to be poor indicators for the actual genotype, due to repeats at breakpoints. A DOC normalized to the average coverage (abbreviated as DRA) between ≥ 0.2 and <0.8, or >1.2 and ≤1.75 were defined heterozygous (0/1) and a DRA <0.2 or >1.75 as homozygous (1/1). In case multiple samples were processed, CNVnator calls were first merged across samples requiring the same event type (gain or loss), and a reciprocal overlap of at least 70% or the non-overlapping parts of each call to be at most 200 bases. For merged calls, the average start and end coordinates, and call properties were calculated. CNVnator calls were then merged to Lumpy calls using the same merging criteria, preserving the more precise start and end coordinates of Lumpy’s SV calls that are based on SRs and PEs.

QC metrics from the input and output data of *ClinSV* are automatically collected for each sample and compared to the MGRB control cohort. The deviation from the control cohort’s average is reported as z-score. Samples passed QC if most input metrics were within an absolute z-score of 2.

To annotate SV with features, such as DGV variants or genes, features were stored in BED, VCF or GFF format, indexed using tabix and queried using the Perl module tabix (https://github.com/samtools/tabix/tree/master/perl, v0.2.0).

### Depth of coverage annotation

For annotating SV calls with DOC, bam sequence alignment files were converted to BigWig [38] format and queried using the Perl module Bio-BigFile-1.07. The DOC of a variant was normalized by the DOC of its flanking region (DRF: DOC ratio flanking) or the genome average (DRA). DRF was calculated by dividing the DOC within the variant by the average of both flanking regions of the same length as the variant. The DOC for the DRF and DRA computation was determined if possible from genomic regions with an average read mapping quality of >=50 (Phred scale; max 60 for BWA MEM), unless less than one third of the variant length fulfilled this criterion, in which case no mapping quality cut-off was applied.

The average genome read coverage was estimated from a 10 Mb window (20,000,001 to 30,000,000) in each autosome, which was chosen to not overlap any centromeric or telomeric regions. The autosomal read coverage was set to the median coverage of the 10Mb region across all autosomes. For the Y chromosome the coverage was estimated from the two large unique regions (6,641,419-10,079,253 and 13,800,704-23,668,908) and for X the entire chromosome was used, to be robust against large copy number changes. To further increase the robustness of the sex chromosome coverage estimates, their coverage was rounded in fractions of 0.5 of the autosomal coverage.

SV calls from Lumpy and CNVnator were classified as CNV (CNV=1) if DRA or DRF was <0.8 or >1.2. The population coverage standard deviation was computed from DRA of 500 healthy control samples in adjacent intervals of 1 kb. Most of the human genome (85%) had a standard deviation of <0.15 (Suppl. Fig. 1).

### Determining SV population variant allele frequencies

Five population allele frequency (PAF) estimates were calculated. Three were derived from a control cohort of 500 healthy Australian individuals as part of the Medical Genome Reference Bank; [36,37], one from gnomAD [31] and one from the 1000 Genomes Project [12] (PAF1KG, Suppl. Fig. 2).

One PAF derived from the MGRB cohort was based on the sum of DP and SR counts (referred to as PAFSU), another was based on the normalized read depth (referred to as PAFDRA) and the third was based on the actual Pass and High confidence variants (referred to as PAFV). MGRB variants were joint-called in batches of 15 samples. PAFSU was calculated in order to obtain an abundance estimate of variants with few DPs and SRs close to or below the detection threshold (Suppl. Fig. 3). PAFSU is computed from the sum of DP and SR supporting a variant (SU) and the corresponding sum of DP and SR in the control cohort (SUC) averaged by the number of control samples (NCS, PAFSU=SUC / SU / NCS). E.g. if a variant has in total 10 supporting DPs and SRs, and if in the control cohort (n=500) there are in total 200 matching DPs and SRs, then PAFSU is 4%. Control sample’s DPs and SRs had to be located within 1,000 bases flanking each breakpoint and orientated corresponding to the SV type (DEL, DUP, INV). PAFDRA was calculated to obtain an abundance estimate for CNV in regions of segmental duplications lacking DPs and SRs often having imprecise boundaries or being fragmented (Suppl. Fig. 4). PAFDRA is obtained by counting the number of samples that show the same copy number change (gain or loss) over at least 90% of the CNV. To speed up the comparison process the DRA of the control samples was summarized in adjacent intervals of 1,000 bases. This summarized coverage was also used to compute the population coverage standard deviation. When determining concordant variants for assigning PAF1KG and PAFV, the variant type (DEL, DUP, INV) including the CNV state (CNV=1 for gain or loss, CNV=0 for copy number-neutral calls) had to match. For copy number-neutral SV, each breakpoint position could deviate at most 1,000 bases. For CNV at least 20% of the reference variant length and 90% of *ClinSV* variant length had to match. For *ClinSV* variants smaller than 1,000 bases at least 70% of their length had to match.

### Variant prioritization

A variant was marked as common if any of the PAF values (PAFV, PAFSU, PAFDRA or PAF1KG) were >0.01; remaining variants were declared rare (RARE=1). Variants affecting genes present in the provided gene list will contain the corresponding gene name in column CANDG. When jointly analyzing multiple individuals and providing pedigree information in ped format, column IA and IUA show how often a variant was present among affected and un-affected individuals, respectively.

### Additional SV annotation

*ClinSV* variants were also annotated with SV from DGV [30], using the same variant matching criteria as for PAF1KG. Additional SV annotation included the variant’s GC content and the sequence compression ratio (CR). The latter is a measure for the repetitiveness of a sequence and used to indicate tandem repeats. For this the compressed sequence length (using perl Compress::Zlib) is divided by the actual sequence length. Accordingly, a tandem repeat region has a lower CR than a complex sequence. SV were also annotated with previously published segmental duplications [39] obtained from genome.ucsc.edu. In cases where more than one segmental duplication overlapped with an SV, the best matching was selected, which was defined as the one with the highest sum of overlap length (in % of variant length) and sequence similarity (in %, sum of both). For annotating SV with genes they affected, ENSEMBL genes release 75 for GRCh37 [40] excluding pseudogenes was used. Gene to disease phenotype associations were obtained from OMIM, Orphanet and DDG2P via ENSEMBL Biomart GRCh37 [40].

### Analytical performance -sensitivity using ClinVar

Pathogenic and likely pathogenic ClinVar variants between 50 bases and 1 Mb were obtained from the UCSC table browser May 2020. Variants were inserted *in silico* into two GRCh37 reference sequences, one for the copy number losses including variants marked as deletions and one for gains including duplications. Variants were required to not overlap and respecting a 5kb flanking region, prioritizing smaller over larger variants. Excluding 4 variants in masked regions of the genome build GRCh37, we obtained 2003 deletions and 467 duplications. Illumina paired reads were generated *in silico* from each reference using ART [41] to an average coverage of 15x depth and combined with an additional 15x depth of the unmodified reference excluding chromosome X and Y, resulting in two 22xy genomes heterozygous for autosomal variants and homozygous for variants on X and Y. Variants were detected with *ClinSV* using default settings. Variants were considered concordant if their breakpoints were within 1,000 bases or if a reciprocal overlap of at least 80% of the variant length was observed, allowing multiple matching parts given all parts were HIGH or PASS.

### Analytical performance -sensitivity using GIAB

We assessed the sensitivity of *ClinSV* to detect CNVs using reference cell lines and gold standard variant callsets developed by the GIAB consortium for NA12878 (aka GIAB1) [33] and an Ashkenazi parent-child trio (aka GIAB2) [33]. We used independent long-read PacBio sequencing data [33] to filter out 12 CNV deletions > 500bp from GIAB1 that lacked any support from both our short-reads and PacBio long-read sequencing data. The more recent GIAB2 was supplied without read depth information, so duplications were distinguished from insertions based on the uniqueness of the inserted sequence as flagged by GIAB. We sequenced NA12878 reference material nine times and the child from the Ashkenzi trio once. SV were detected for *ClinSV*, Lumpy, CNVnator, Delly2 (v0.7.6) and Manta (v-1.1.1) with default parameters, from a single replicate of NA12878 processed as described above. Here we defined sensitivity as the fraction of true positives identified from the set of gold standard calls: TP/(TP+FN). Concordance criteria see sensitivity using ClinVar.

### Analytical performance – false positive rate using PacBio

The same NA12878 Illumina short-read data was used as input to call High and Pass variants for *ClinSV*, Lumpy, CNVnator, Delly and Manta. For each caller 50 deletion, 50 duplications greater and smaller than 500 bases (200 variants per caller) were randomly selected and compared to the GIAB SV calls (deletions only), PacBio NA12878 SV calls and PacBio read alignments. PacBio SV calls were obtained from the Genome in a Bottle (GIAB) resource web page (http://jimb.stanford.edu/giab-resources/ files last modified April 2015). These SV calls from GIAB were based on three different variant detection methods: PBHoney [42], a custom pipeline on assembled sequences (unpublished), and the Chaisson et al. methodology [43]. In addition we called SVs with Sniffles v1.0.3, after realigning the PacBio reads using NGMLR v0.2.1 [44]. SV calls obtained from *ClinSV* or the GIAB gold standard [33] were considered valid, if they were present in at least one of the above PacBio variant call sets or if clearly visible in PacBio read alignment or read coverage profile, i.e. deletions and duplications smaller than PacBio read length were required to have SR support and those larger DOC support.

### Analytical performance – false positive rate using MLPA

MLPA was used as orthogonal confirmation method to assess the false positive rate for *ClinSV* and to validate candidate CNV during the ‘application to clinical genetic testing’ section below. MLPA assay setup was performed as per standard protocols which also included 4 control probes C1, C2 (binding to exons of EP300 and CREBBP), NR0B1 (binding chrX) and SRY (binding chrY) per batch. MLPA design was attempted for 2 probes for each variant as per standard protocols to allow redundancy in case of probe failure. CNVs not confirmed by MLPA were considered false positive calls.

### Analytical performance – Reproducibility

Pass and High confidence *ClinSV* calls from nine NA12878 replicates were compared in an all versus all comparison. SV concordance were as for the GIAB1 sensitivity analysis.

### Analytical performance – Comparison to microarrays

DNA samples of 11 patients from the molecular cytogenetics laboratory at South Eastern Area Laboratory, Sydney Australia (cohort A) were analyzed on the Agilent 8×60k ISCA v2 design array (031746). The design of this array consists of 59,059 distinct target probes, and 3,886 control probes. The array was semi-targeted, with 18,851 densely tiled probes located in cytogenetically relevant disease regions. Probes located in the “backbone” regions had a lower tiled density. Overall median probe spacing was 60 kb however this will be higher in regions of high probe density. 10/11 samples were run in a duplicate dye swap experiment design (one replicate sample labeled with Cyanine-3-dUTP, one replicate sample labeled with Cyanine-5-dUTP). The remaining sample was run in singlicate.

DNA samples of 17 patients from the molecular cytogenetics laboratory at the Children’s Hospital Westmead, Sydney Australia (cohort B) were analyzed on the Agilent 2×400 kb CGH array. This array comprises of 411,056 distinct biological probes and 5,232 control probes with a median probe spacing of 5,315bp. Raw MA files were analyzed using Agilent CytoGenomics software v4.0.2.21 with the default calling algorithm (Default Analysis Method – CGH v2).

All MA calls were manually reviewed and classified as Low, Medium or High confidence by highly experienced cytogeneticists (Suppl. Table 1). Low confidence calls were excluded from further analysis. Classification of confidence was based on minimum log2 deviation, minimum probes, frequency in the cohort B (high frequency indicative of potential MA artifact or copy number polymorphism) and the call being visually consistent with a copy number change (consistency of vertical scatter of probes, location within a chromosome). MA calls were lifted over from hg18 to the GRCh37 reference genome build. High and Pass confidence calls from *ClinSV* were compared to the MA calls, requiring at least 50% reciprocal overlap. Each discordant variant was visualized in a genome browser. CNV with an overlap of 0–50% were reclassified as concordant imprecise.

### Flanking repeat characterization

We sequenced NA12878 by WGS (library FR05812606), ran *ClinSV*, and for each Pass CNV (n=4,634), the sequences surrounding each breakpoint were retrieved, including up-to 2 kb up and downstream. If the CNV was smaller than 4,000 bases, the inner ends were trimmed to assure non-overlapping sequence pairs. For each CNV, the pair of breakpoint sequences were aligned to each other using NCBI blastn v2.3.0+ [45] (default settings). Repeats at the breakpoint obtained from the sequence alignment were required to cross each breakpoint by at least one base. To classify the repeats, Tandem Repeats finder (v4.0.9) [46] was applied to the upstream SV repeat using default settings. SV repeats for which no tandem repeat was identified were subsequently intersected with GRCh37 RepeatMasker annotation [47] using Bedtools (v2.25.0) [48].

### Application in clinical genetic testing

Patient blood samples were submitted for diagnostic testing to Genome.One, a clinically accredited laboratory in Sydney, Australia from March 2018 to December 2019. DNA samples were extracted using standard protocols and subject to WGS as described above, although some samples used KAPA Hyper PCR-free library preparation kits. Following WGS and *ClinSV* analysis, CNVs for each patient were filtered by PAF1KG <2% and PAFMGRB <2% and restricted to High or Pass confidence. The analysis was mostly restricted to disease specific curated gene lists, or virtual gene lists consistent with the patient’s presenting phenotype; CNVs were filtered to only those overlapping these genes. Where whole genome analysis was requested, analysis was limited to currently described disease associated genes in OMIM. Analysis was restricted to genic regions +/-200bp.

The CNV interpretation considered the overlap between the patient phenotype and that described for the disease associated with the deleted or duplicated gene, inheritance model of the disease, zygosity of the CNV and any other SNVs or indels detected by standard pipelines. Classification of CNVs was based on a combination of the ACMG standards and guidelines for the interpretation of sequence variants[49] and the ACMG standards and guidelines for interpretation and reporting of postnatal constitutional copy number variants [50]. Variants were described using HGVS v15.11.

### Resolution of WGS CNV by clinical microarray

The major Australian accredited clinical diagnostic laboratories were surveyed to identify commonly used array designs for routine diagnosis of genetic diseases and the minimum number of probes required to call a CNV. Five array designs were used across seven laboratories (Suppl. Table 2). Array designs were downloaded from vendor websites and where required converted to bed file format. CNVs detected by WGS were converted to BED file format and bedtools intersect was used on each array design to extract the probes within the breakpoints of each CNV. Each CNV was deemed undetectable for an array design if it contained fewer than the minimum required probe number for a call within the breakpoints of the CNV.

## Results

### ClinSV

To identify clinically relevant CNVs and copy number-neutral SVs from Illumina short-read WGS data, we developed *ClinSV*, which is a computer-assisted analysis, annotation and visualization framework, summarized in Fig. 1c, and described here.

### Variant Identification

CNVs larger than 50 bp were detected using three evidence types, DP, SR and changes to DOC (Fig. 1a), while copy number-neutral SVs were detected using just DP and SR. SR generally allow detection of genomic breakpoints with base-pair resolution (Fig. 1b). *ClinSV* integrates CNV calls from CNVnator [14], which uses evidence only from DOC, and Lumpy [15], which uses evidence only from DP and SR. *ClinSV* then determines the level of SR and DP support for each candidate CNV from CNVnator and computes the change in DOC for each candidate CNV from Lumpy before merging overlapping calls (see methods). This supports the identification of higher confidence CNV calls with multiple lines of support.

### Variant quality

The merged variant calls are assigned an automated confidence category, (High, Pass, Low) based on the amount of supporting evidence and CNV size (Suppl. Table 3). Based on 30–40x WGS from Illumina HiSeq X, we defined the following criteria: High confidence CNVs are either large (>100 kb), or >10 kb with confidently mapped reads (i.e. average mapping quality >55 across the entire variant). These criteria reduce false positives due to segmental duplications (Suppl. Fig. 5 and 6). Pass CNVs are either >10 kb, or ≤10 kb with at least two supporting split or discordant read pairs. High confidence copy number-neutral SVs need at least 10 split or spanning read support, and at least one from each source, whereas Pass copy number-neutral SVs need at least 6 supporting reads. All other CNVs or SVs calls are flagged as Low confidence.

### Quality attributes

Quality attributes of each candidate SV are reported, including the average mapping quality at breakpoints and across the SV, the average %GC content, the sequence complexity via its compression ratio (see methods), and overlapping segmental duplications [39]. Low mapping quality or extreme %GC indicate possible false positives due to mapping artifacts or GC-coverage bias. CNVs with small sequence compression ratios usually represent variants in short tandem repeats, which often have reduced DP and SR evidence from short-read sequencing (see below).

### Annotation

SVs are annotated with variant allele frequencies from 500 healthy controls (see below), overlapping genes, as well as their known disorders from OMIM [51], and phenotypes as Human Phenotype Ontology (HPO) terms [52] from Orphanet [53].

### Variant filtration

*ClinSV* identifies 4,730±190 CNVs (mean ± stdev), and 1,100±150 balanced SVs per germline 30–40x genome. Whilst *ClinSV* does not systematically classify the copy number-neutral SVs into subtypes, the 1,100 SV are mostly mobile element insertions (MEIs), ∼50 inversions, and almost no translocations; these findings are consistent with previous genome-wide estimates from short-read sequencing data [15]. By restricting to rare, gene-affecting variants, *ClinSV* obtains on average 11 CNVs and 5 copy number-neutral SVs per individual. *ClinSV* also allows joint variant calling of multiple samples, so if pedigree information is provided, variants that are associated with only affected individuals can be readily identified. If a set of candidate genes is supplied, this set is further reduced; for example, in ∼100 familial dilated cardiomyopathy genes, on average only 0.5 rare SVs were identified per patient [10]. Finally, *ClinSV* annotates variants using OMIM and Orphanet terms which can be useful to prioritize variants in patients’ with challenging phenotypes.

### ClinSV output files

*ClinSV* produces a text file of all annotated CNVs and SVs, and a smaller file with just the rare, gene-affecting CNVs and SVs (Suppl. Data File 2, Suppl. Table 4). To assess whether the input data and *ClinSV* results are within expectations, a comprehensive QC report is also generated, including metrics on the fragment length distribution, chimeric read counts, read coverage variability, genome wide DOC, number of called variants stratified by caller, variant type and properties including the frequency of gene-affecting and rare SVs (Suppl. Data File 3). Additionally, when run on the NA12878 cell line, a comprehensive analytical validation report summarizing sensitivity and reproducibility is produced. Metrics outside 2-standard deviations from the average value determined from 500 healthy control samples are flagged for review.

### Visualization framework

Manually reviewing candidate variant calls remains common practice in most research and clinical laboratories and is particularly important for SVs. *ClinSV* creates an IGV session file, which loads 11 tracks allowing the assessment of variant validity and potential clinical relevance (Fig. 2). We have provided recommended criteria to evaluate whether each candidate variant passes, needs further investigation, or is a likely false positive (Suppl. Table 5).

**Figure 2.**
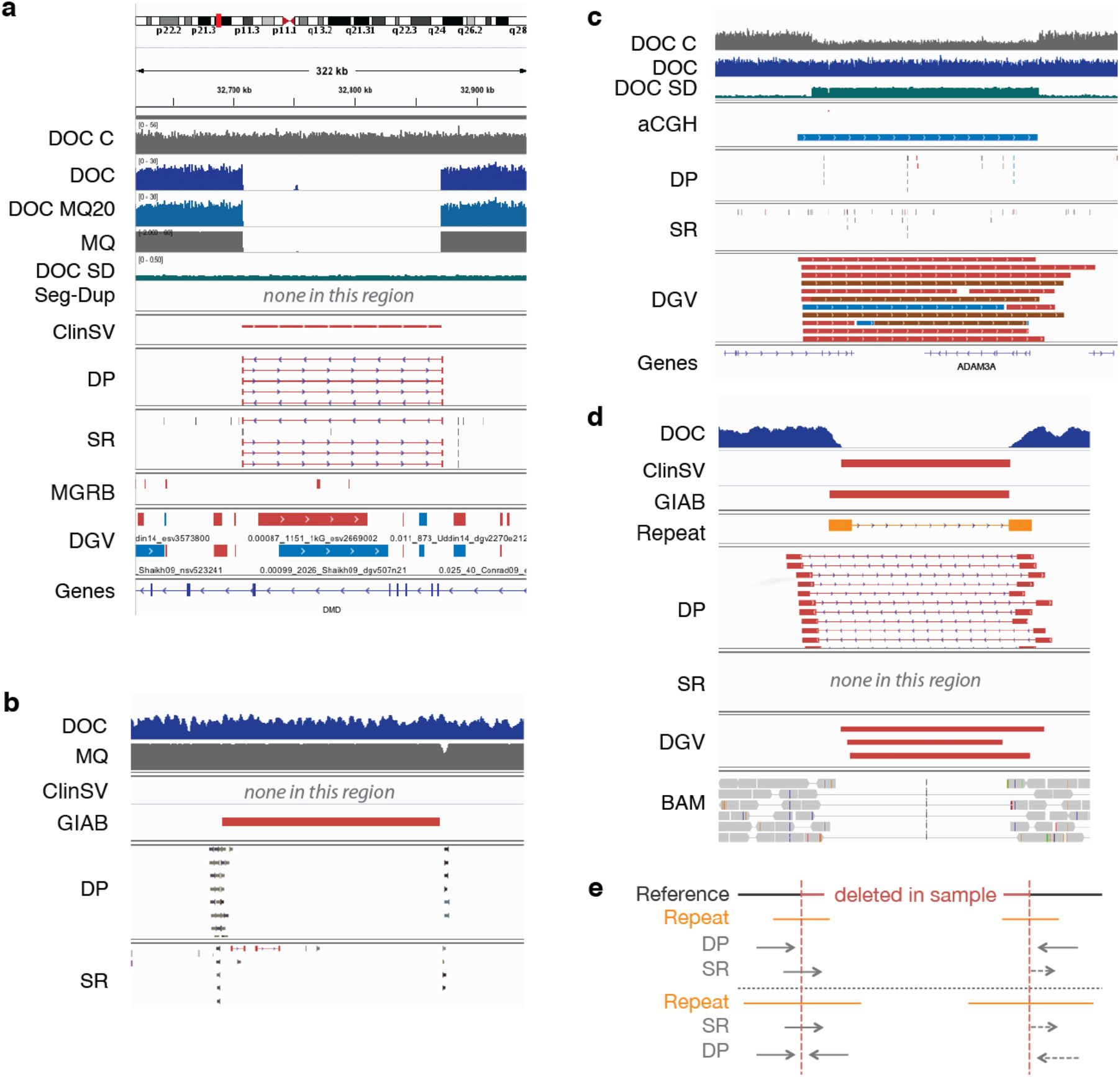
Properties and supporting evidence for accurate structural variant identification. **A:** A pathogenic homozygous deletion in the Dystrophin gene. *ClinSV*’s default annotation tracks are: depth of coverage in an NA12878 control (DOC C), all reads in the sample (DOC), or reads with mapping quality >=20 (DOC MQ20), the average mapping quality of aligned reads from the sample (MQ), coverage standard deviation from 500 controls (DOC SD), segmental duplications [21] (Seg-Dup), annotated *ClinSV* calls (*ClinSV*), discordant pairs (DP), split reads (SR), *ClinSV* variant calls from 500 controls (MGRB), variants from the Database of Genomic Variants (DGV), and RefSeq genes (Genes). **B**: A representative false positive CNV deletion identified by GIAB: the 5’ breakpoint is a mobile element insertion (MEI), the 3’ breakpoint is located in a repeat as suggested by the lower read mapping quality, and there is no intervening drop in coverage or mapping quality. **C:** A representative false positive duplication called by aCGH not supported by WGS. The region shown is deleted in 44% of the samples in the 1000 Genome Project and coverage is frequently altered in controls (see DOC SD), but not in the actual sample (see DOC). **D:** A homozygous deletion identified by *ClinSV* with reduced DOC, lots of DP support, but no SR support. The CNV is flanked by a pair of repeats. **E:** A model demonstrating how CNV surrounded by repeats can reduce DP and SR support. For repeats greater then read length but smaller than paired-end fragment size only SR support is reduced (top, as in panel d), for repeats greater than the fragment size, both SR and DP support are reduced (bottom). The dashed arrows indicate the expected mapping location of DPs and SRs if the genomic region was not repetitive.

### Orthogonal validation

Variants passing manual review were orthogonally confirmed using either multiplex ligation-dependent probe amplification (MLPA) or Sanger sequencing.

### Scope

*ClinSV* identifies CNVs >50bp in length, and copy-number neutral SV if breakpoints are located in unique regions of the genome. ClinSV has been developed predominantly to support rare disease diagnosis workflows within the nuclear genome, but can identify SV in the mitochondrial genome, mosaic variants and tumour genomes given enough supporting evidence, which will be evaluated more formally elsewhere.

### Performance

Starting from a coordinate-sorted 30–40x depth BAM file, *ClinSV*’s runtime is 8 hours on a single compute node (48 GB RAM, 16 CPUs, Suppl. Table 6). This fast performance is facilitated by rapid extraction and processing of SR and DP reads, and by running a parallelized version of CNVnator from SpeedSeq [21].

### Allele frequencies from healthy individuals

To assist with the prioritization of rare SVs, we ran *ClinSV* on a cohort of 500 healthy individuals, sequenced with the same technology (i.e. 30–40x depth WGS from a single-lane of Illumina HiSeq X) and TruSeq Nano HT v2.5 library preparation chemistry. These healthy individuals were sequenced as part of the Medical Genome Reference Bank (MGRB [36,37]). We identified 922,161 High, 1,998,221 Pass and 1,991,316 Low quality non-unique SVs across the cohort. We also evaluated the raw supporting read evidence, as highly repetitive parts of the genome may have highly variable or poor coverage, or have few split or spanning reads. We considered the raw evidence from SR, DP, and DOC, as well as final SV calls to derive three population allele frequency (PAF) measures from MGRB (see Methods). The underlying database or raw evidence data is included in the *ClinSV* distribution and the derived PAF capture common complex variants or mapping artifacts that often do not result into variant calls. In addition we annotate variants with population allele frequencies from gnomAD [31] and the 1,000 genomes project [12]. Variants are labeled as rare if none of the computed PAFs were greater than 1%, reducing the number of candidate variants to inspect >100-fold.

### Analytical validation

We assessed the analytical performance, including sensitivity, false positive rate and reproducibility of *ClinSV*, by comparing to pathogenic ClinVar variants, Genome in a Bottle (GIAB) consortium gold standard reference materials [33,35], long-read sequencing, MLPA and clinical MA. Each technology and derived call-set had its own strengths and limitations in representing the different SV types, sizes and pathogenic variants.

### CNV sensitivity

The sensitivity was assessed using 2,470 pathogenic deletions and duplications replicated in-silico and two GIAB standards derived from cell lines of healthy individuals (NA12878, NA24385).

In order to assess performance of ClinSV on structural variants that are clinically relevant, we identified 2,470 pathogenic and likely pathogenic CNVs from the ClinVar database and generated a simulated 30x-depth WGS dataset, where each variant was simulated as affecting one allele (Methods). Among the CNVs larger than 10kb, all duplications (281/281) and all but two deletions (674/676) were detected with *ClinSV* (99.8% sensitivity). Both missed deletions were partially identified, where one was interrupted by a segmental duplication and the other was split in two and lacked flanking SR and DP support. Smaller than 10kb the sensitivity was 83.4–96.5% for the deletions and 41.1–85.7% for the duplications (Table 1).

**Table 1.** ClinVar pathogenic CNV sensitivity analysis. The sensitivity of ClinSV was assessed using simulated WGS data representing 2,470 pathogenic or likely pathogenic CNVs.

| Size range | Deletions |  |  | Duplications |  |  |
| --- | --- | --- | --- | --- | --- | --- |
|  | # Variants | Sensitivity | # Missed | # Variants | Sensitivity | # Missed |
| 0-500 | 753 | 83.4% | 125 | 95 | 41.1% | 56 |
| 500-1k | 104 | 91.3% | 9 | 9 | 77.8% | 2 |
| 1k-5k | 328 | 94.5% | 18 | 47 | 89.4% | 5 |
| 5k-10k | 141 | 96.5% | 5 | 35 | 85.7% | 5 |
| 10k-50k | 278 | 99.6% | 1 | 79 | 100.0% | 0 |
| 50k-100k | 123 | 100.0% | 0 | 28 | 100.0% | 0 |
| 100k-500k | 207 | 100.0% | 0 | 110 | 100.0% | 0 |
| 500k-5M | 69 | 98.6% | 1 | 64 | 100.0% | 0 |
| Total | 2003 | 92.0% | 159 | 467 | 85.4% | 68 |

We also investigated ClinSV sensitivity using two GIAB gold standard datasets (aka GIAB1, GIAB2, see Methods), where again the sensitivity also generally increased with size, reaching 97–100% for variants >10kb (Fig. 3a, Suppl. Table 7–9). The overall sensitivity for detecting deletions was 88% for GIAB1 and 72% from GIAB2. Missed variants had little or no DP/SR evidence due to flanking repeats (Suppl. Fig. 7 and see below) or had conflicting DOC increase due to poor read alignment within the deleted repeat (Suppl. Fig. 8). GIAB1 was primarily based on Illumina short reads, whereas GIAB2 also included large amounts of long-read data, known to detect more than twice as many deletions and duplications in tandem repeats [43]. In comparison to the two other popular SV callers Delly2 [16], Manta [17], ClinSV performed better for deletions

**Figure 3.**
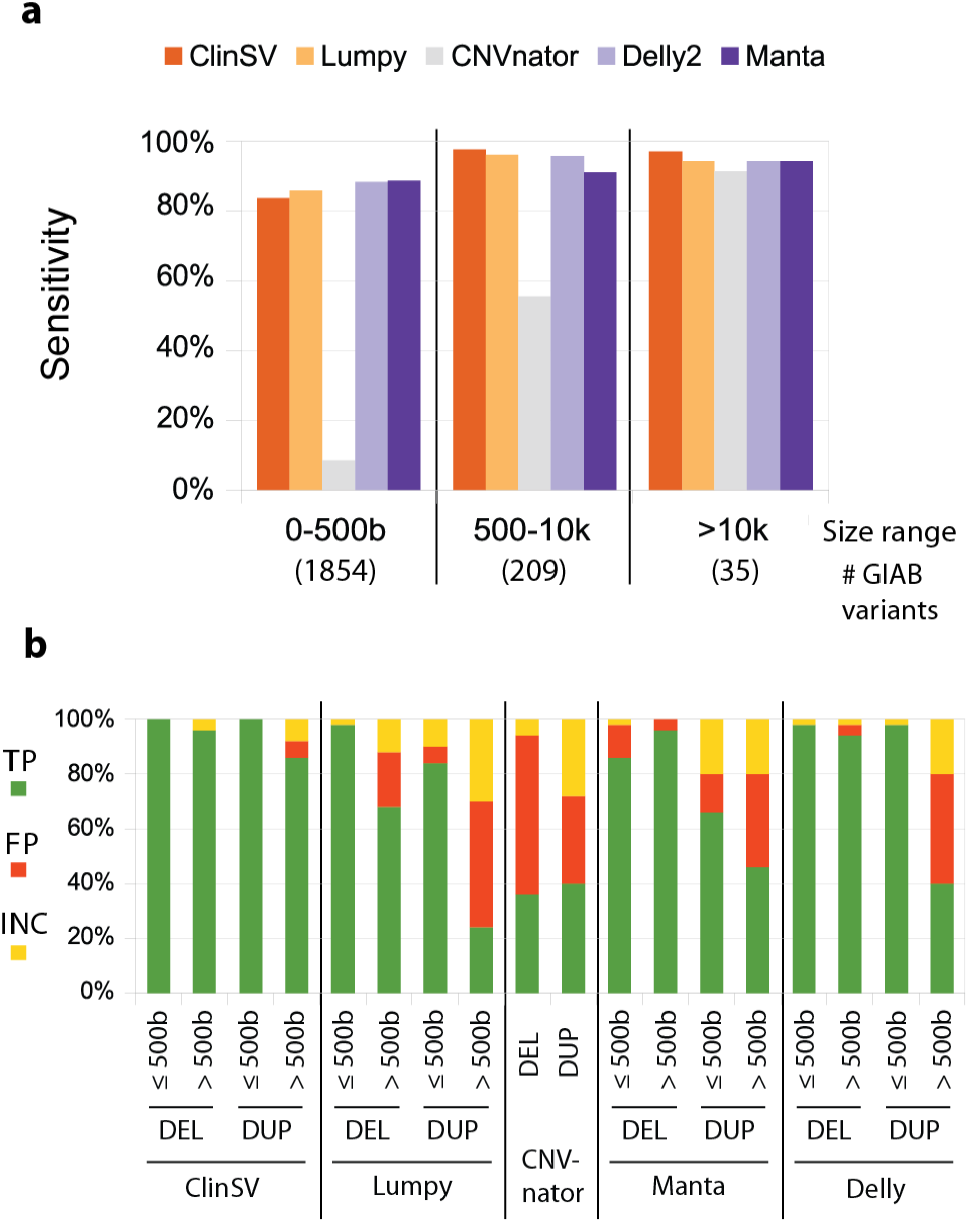
Analytical performance of *ClinSV*. We sequenced the NA12878 control cell line using WGS and compared the performance of a number of popular CNV or SV detection methods. **A:** The sensitivity of different methods to identify 2,664 CNV deletions from the Genome in a Bottle (GIAB1) gold standard call set over different size ranges. The number of deletions per size range are indicated along the bottom. **B:** We selected deletions and duplications larger, or smaller than 500 bp (n=50 each; total n=200) from each of five different methods. CNVnator had few calls < 500 bp, so we combined all calls into one category. We assessed whether each call was a true positive (TP), false positive (FP) or inconclusive by comparing to long-read PacBio data.

>500 bases but slightly worse for duplications (Fig. 3b, Suppl. Tables 7–9). The overall sensitivity to detect presumably benign duplications <10kb, as represented by the GIAB2 standard was poor for all tested short-read based variant callers at 1.4–43.8% (Suppl. Table 9).

### CNV false positive rate via PacBio

We evaluated the false positive rate (FPR) of CNVs identified by *ClinSV* and other popular callers using the NA12878 cell line and CNV calls made using PacBio long-reads [44]. Deletions and duplications smaller or larger than 500 bp were randomly selected (50 each, total 200 per variant caller) and categorized into three bins: present in control data (true positives), absent (false positives), and inconclusive. The FPR is shown as a range, with the smaller number corresponding to the proportion of false positive calls to the sum of true and false positives, whereas the larger more conservative number counts inconclusive calls as false positives. The FPR was lowest for *ClinSV* (1.5–4.5%), followed by Manta (11.8–17.5%), Delly2 (17.9– 26.5%), Lumpy (20.8–31.5%) and CNVnator (54.2–62.0%; Fig. 3b, Suppl. Table 10). 100% of the High confidence *ClinSV* deletions were true positives (n=71), thus High confidence calls had a FPR of 0%. The visualization, annotation, and population allele frequency data, which support effective manual variant review, are critical elements of *ClinSV*, contributing to its lower FPR compared to the other four callers.

### CNV false positive rate via MLPA

The FPR of CNV calling from WGS using *ClinSV* was also assessed using custom MLPA which is a targeted assay for the detection of CNVs. From six patients, we selected 29 High and Pass CNV candidates that passed manual review, overlapped genes (+/-200bp) and were rare (PAF <2%). Custom MLPA assay design was successful for 26 of these CNV candidates, all of which confirmed the presence of the CNV (FPR 0%, Suppl. Table 11). If we conservatively count the three CNV for which an MLPA assay could not be developed as false positives, then the FPR of *ClinSV* is 10.3%. Furthermore, we selected 10 gene-overlapping, rare CNV candidates that failed manual review (see visualization framework). Custom MLPA assay design was successful for three of the ten, with the assay confirming all three variants as true positives. This highlights that even some variants that fail manual review – an important component of the *ClinSV* procedure – may still be true positives and that these variants, if located in relevant disease genes may warrant further investigation.

### Reproducibility

We sequenced nine NA12878 replicates using three library batches, two different technicians, two sequencing runs, two sequencer sides, and two different lane positions (Suppl. Table 12). In an all-versus-all comparison, on average 99.1% of High confidence CNVs and 98.7% of High confidence SVs were detected in technical replicates (Table 2). When combining Pass and High confidence CNVs, the reproducibility was 85.0% and 83.9% for all SVs. A pairwise comparison of all replicates revealed similarly high concordance in all cases (Suppl. Fig. 9).

**Table 2.** Reproducibility of *ClinSV*. Eleven replicates of NA12878 were sequenced using 30–40x WGS, and the consistency of CNV and SV calls identified by *ClinSV* was assessed.

| SVs/CNVs | Confidence | Average | Stdev | Min | Max |
| --- | --- | --- | --- | --- | --- |
| CNVs | High | 99.1 | 0.3 | 98.5 | 99.7 |
| CNVs | High & Pass | 85.0 | 1.9 | 81.3 | 88.8 |
| All SVs | High | 98.7 | 0.4 | 97.5 | 99.4 |
| All SVs | High & Pass | 83.9 | 2.6 | 78.1 | 89.1 |

### Concordance with patient microarrays

We compared CNVs detected by *ClinSV* from WGS to CNVs detected by clinically accredited MA. A total of 28 patients from two patient cohorts underwent WGS (Illumina HiSeq X 30–40x) and MA aCGH. One cohort of 11 samples consisted of patients with 11 pathogenic CNVs identified on Agilent 60k CGH arrays (referred to as cohort A). The other cohort of 17 samples analyzed on Agilent 400k CGH arrays (referred to as cohort B) had a total 284 CNV calls, none of which were classified as pathogenic.

Of the pathogenic CNVs in cohort A, 100% (11/11) were identified using *ClinSV* (Table 3). Looking in more detail, CNVnator identified 9/11 CNVs, Lumpy identified 4/11 CNVs, and two sex chromosome aneuploidy events were identified only by *ClinSV*’s automated sex chromosome ploidy algorithm. Thus, even for the detection of large CNV within the size range of aCGH, it is critical to integrate evidence from DOC, DP, SR, and chromosome-wide coverage analysis (Suppl. Data File 3, Section 3).

**Table 3.** Concordance of *ClinSV* with pathogenic variants from aCGH. Comparison of pathogenic CNVs identified by aCGH and WGS using *ClinSV* on matched DNA from the same patients. Eleven patients with known pathogenic variants were compared. Genomic coordinates are listed compared to the GRCh37 reference genome build. Sample A4 had a large CNV identified by aCGH, which was disrupted by a 7.3 kb diploid region in the middle.

| Sample | aCGH calls | WGS calls |
| --- | --- | --- |
| A1 | 22q11.21(18,894,835-21,505,417)x1 | del(22)(q11.21q11.21)<br>chr22:g.18,873,701_21,466,500del |
| A2 | 22q11.21(18,894,835-21,505,417)x3 | dup(22)(q11.21q11.21)<br>chr22:g.19,025,201_21,502,400dup |
| A3 | 15q11.2q13.1(23,656,936-28,559,402)x1 | del(15)(q11.2q13.1)<br>chr15:g.23,679,001_28,659,900del |
| A4 | 15q11.2q13.3(22,765,628-32,418,879)x3-4 | dup(15)(q11.2q13.2)<br>chr15:g.22,727,651_30,670,600dup,<br>dup(15)(q13.2q13.3)<br>chr15:g.30,677,901_32,679,700dup |
| A5 | (X)x1 | del(X)(p22.33q28) chrX:g.pter_cen_qterdel |
| A6 | (X)x2,(Y)x1 | dup(X)(p22.33q28) chrX:g.pter_cen_qterdup |
| A7 | 4p16.3(1,832,643-1,994,922)x1 | del(4)(p16.3p16.3)<br>chr4:g.1,830,325_1,996,237del |
| A8 | 16p11.2(29,849,168-30,190,568)x1 | del(16)(p11.2p11.2)<br>chr16:g.29,560,701_30,200,100del |
| A9 | Xp22.33p22.31(60,701-8,392,011)x1,<br>Yq11.221q12(16,188,682-59,335,913)x1 | der(X)t(X;Y)(p22.31;q11.221)<br>g.[chrX:pter_8,434,700<br>delinschrY:16,097,288_qterinv] |
| A10 | Xp21.1(32,714,410-32,868,014)x0 | del(X)(p21.1p21.1)<br>chrX:g.32,707,584_32,871,077del |
| A11 | 15q11.2(22,765,628-23,077,909)x3 | dup(15)(q11.2q11.2)<br>chr15:g.22,727,651_23,377,700dup |

We compared the total set of MA calls from either clinical laboratory to *ClinSV* calls from WGS and initially found a concordance rate of 62–75% (Suppl. Table 13). We manually inspected all of the n=95 calls found only by aCGH. In cohort B, 64 of the 70 calls found only by aCGH were located in regions of common copy number variation, which often coincided with multi-allelic CNV [54], where the WGS data was clearly diploid (no DOC, SR, DP, in regions with good mapping quality). Since aCGH uses a pool of DNA from controls as a comparator, if a CNV is a common deletion in the general population, then a diploid patient may have an erroneous CNV amplification called (Fig. 2c), and vice versa (Suppl. Fig. 10). Whilst these 64 events represent false positives calls in the MA data, in clinical practice, common copy number variants are usually filtered out, and not reported. In the cohort A, 4/25 discordant calls were in regions with common CNVs, 4/25 were not reproduced on the dye swap, 13/25 had less than four aCGH probes, 3/25 had poor logR intensity deviation from normal (2n) or probe intensities with a high standard deviation (Suppl. Table 13). There was one CNV call made by the 60K aCGH array, of just 528bp with more than 4 probes, for which we find no supporting evidence in the WGS data. Taken together, there was at most 1 *bona fide* extra call made by clinical MA that was not identified by *ClinSV*.

The limited probe density of MA affects the precision of the size of the resulting CNV calls [55]. *ClinSV* on WGS data was able to improve the precision of 38 of the 214 concordant calls from the cohort B, by a total of 960 kb (Suppl. Fig. 11). *ClinSV* also identified additional large CNV that were not found in the MA data. In cohort B, there were on average 193 Pass and High confidence extra CNV per patient > 50 kb identified by *ClinSV*. Of these, 95% were in regions of frequent copy number variation in the population (population allele frequency >5%), and thus potentially deliberately excluded from the MA probe design. The other 5% (average 7 per patient) were in repeat regions (average MQ<34), which could thus be missed by MA due to challenges in designing probes for these regions, or false positives in the WGS data.

### Limitations of short-read based CNV detection

Despite all the improvements of short-read WGS based CNV detection over MA, short-reads have inherent limitations which can hamper CNV detection in some parts of the genome. We investigated why some CNV candidates lacked supporting SR and/or DP evidence (Fig. 2d, Suppl. Fig. 12, 13) and found that many had pairs of near identical repeats on either side of the breakpoints. We hypothesized that in this scenario, paired-end read alignment algorithms would prefer to map the read pairs closer together than flanking the CNV (Fig. 2e). In NA12878, 13% (625/4,634) of CNVs had flanking local repeat structures. The repeats ranged between 30–4,000 bp and mostly consisted of tandem repeats (TR) (75%) followed by SINE (13%) and LINE (6%) elements (Suppl. Fig. 14, 15). The average number of SR, DP and repeat length generally decreased with increasing sequence identity (Suppl. Fig. 15c).

### Application to clinical genetic testing

Collectively, these data demonstrated that *ClinSV* could identify CNVs from 30–40x depth Illumina WGS data with high reproducibility, sensitivity and low false positive rates. Furthermore, *ClinSV* enables the identification of copy-number-neutral SVs and overlapping DEL-DUP events [10]. Ultimately these results were used to support the ISO15189 clinical accreditation of *ClinSV* for the analysis of CNVs from WGS data >50 bp, in patients with suspected monogenic conditions.

Following ISO15189 clinical accreditation in March 2018 through the diagnostic genomics laboratory, Genome.One, *ClinSV* was applied to 485 patients with a diverse range of phenotypes including development delay, heart disease, hearing loss, neurological disease and kidney disease. Within this cohort, a total of 23/485 patients (4.7%) had clinically reportable CNV, 100% of which were orthogonally confirmed, 21 using MLPA and two confirmed in alternative laboratories. MLPA or Sanger did not confirm one additional potential pathogenic CNV which had been classified as inconclusive following manual inspection. Of the reported CNVs, 52.2% were pathogenic, 17.4% likely pathogenic and 30.4% were classified as variants of unknown significance (Fig. 4, Suppl. Table 14). The size range of the reported variants was between 500 bases to 1.5 Mb, with 35% being smaller than 10k.

**Figure 4.**
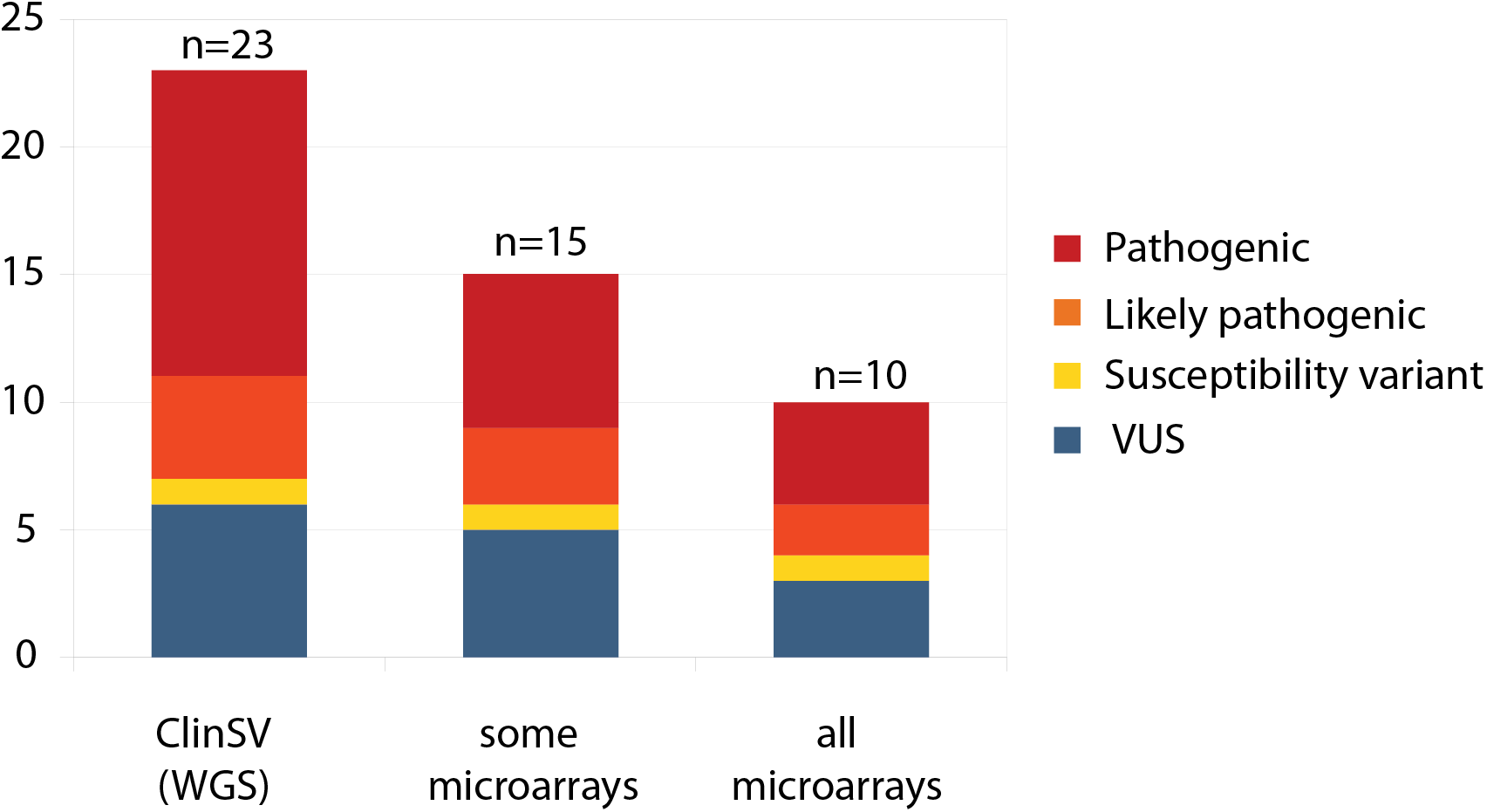
Clinically reported CNVs detected by *ClinSV* compared to callability by clinical microarrays. WGS with ClinSV analysis was applied in a diagnostically accredited laboratory to 485 patients with a diverse range of phenotypes and previous molecular testing. Twenty-three clinically reportable CNVs were identified and subsequently reported to the referring clinician. Only a subset of these would have been identified by all, or some of the microarray platforms available throughout clinical laboratories in Australia.

Four of the reported variants (ID 3, 5, 13, 22) were within genes associated with recessively inherited diseases, where the patients also had an additional rare SNV identified by WGS in the same gene, presumably in a compound heterozygous state: *ERCC5* (MIM133530), *STRC* (MIM606440), *SYNE1* (MIM608441), and *DST* (MIM113810). In these cases, the clinical report included a recommendation to perform family segregation analyses.

Of the clinically reportable variants 8/23 (35%) would not have been detected by any current commercial MA system commonly utilized in Australian clinical diagnostic laboratories and 13/23 (57%) would not have been detected by at least one commercial MA design. Of all pathogenic and likely pathogenic CNVs, 7/16 (44%) would not have been detected by any clinical MA and 10/16 (63%) would not have been detected by at least one clinical MA design (Suppl. Table 14, 15).

The median size of CNV that would be unlikely to be detected by any MA was 3.8kb (range 0.5-50kb). The median size of CNV that were detectable by at least one MA design was 424kb (range 9.3kb-1.5Mb).

## Discussion

Here we have presented *ClinSV*, a platform that enables the accurate detection and interpretation of CNVs and SVs from short-read WGS, with a focus on identifying rare, pathogenic CNVs. To date, most approaches have treated CNV and SV identification as separate analytical challenges; however, as we have shown, there is substantial benefit in combining these approaches.

There are several features that contribute to *ClinSV*’s accuracy. First, it integrates complementary evidence from DOC, DP and SR to obtain a comprehensive set of SVs and CNVs per sample. Secondly, the level of evidence supporting each candidate SV is combined with quality metrics to classify CNVs into High-, Pass-and Low-quality tranches. Excluding Low-quality variants efficiently removed most false positives and kept the sensitivity high relative to other short-read callers. We posit that the poor performance of previous comparisons was due to low quality CNVs affecting the overall quality. Thirdly, the filtration by numerous population allele frequency metrics calculated from a matched sequencing and software pipeline, substantially reduced the number of candidate SVs from 5,800 to an average of only 20 rare gene affecting variants per patient. Fourthly, the integrated visualisation framework adds additional lines of supporting evidence from matched controls that facilitates manual inspection, and further improves the performance of the method. And finally, the comprehensive quality control report is valuable to highlight when the quality of input WGS data is poor; WGS data with variability in sequencing coverage makes CNV identification very challenging and may be caused by degraded DNA or poor-quality sequencing libraries.

By comparing to extensive simulated data, benchmarking and long-read sequencing data now available from the NA12878 and NA24385 cell lines, the analytical performance of *ClinSV* was strong. *ClinSV* detected 99.8% pathogenic ClinVar CNVs >10kb and 85.1% <10kb. The increase in sensitivity for CNV >10kb is due to confident DOC based calls despite the absence of supporting DP and SR, which can occur due to repeats at variant breakpoints.

For presumably benign duplications and deletions >10kb the sensitivity was 97% (DEL GIAB1, DEL/DUP GIAB2) and <10kb ranging between 46–88% depending on the variant type and dataset analyzed. The majority of SVs in healthy individuals are short and in tandem repeats. In contrast, previously reported pathogenic variants have a higher proportion of variants >10kb (Table 1, Suppl. Table 5-7). Long-reads are more sensitive for detecting short insertions and deletions in repeated regions of the genome[43]. Large amounts of such variants are included in the GIAB2 standard.

*ClinSV*’s FPR was 1.5–4.5% overall, substantially outperforming other methods, and demonstrating good performance for CNV duplications, validated by long-read PacBio sequencing data. Variants passing manual inspection and for which an MLPA assay could be designed all validated. Finally, the reproducibility of High confidence CNV calls from *ClinSV* was 99.1%, and 85.0% for High and Pass confidence variants. This reduction in reproducibility of Pass CNV (relative to High CNV) was usually due to variants with small number of supporting reads, close to the detection threshold, which occasionally fell below the automated detection cutoffs. This suggests that higher than 30–40x sequencing depth may improve reproducibility and increase the number of variant calls. The visualization of the region with a suspected variant may reveal additional supporting reads not sufficient to automatically call a variant.

One of the major challenges during the development of *ClinSV* was to obtain high confidence calls to benchmark our methods. Continued efforts by the community to share data [56], benchmark methods [57], and standardize variant call sets [33] are critical for methods development and will play an increasingly important part of translating genomics to the clinic. There are very few validated copy number-neutral SV, or sets of CNV duplications in NA12878 [33] or other healthy controls [12], so comprehensively assessing the performance of *ClinSV* and other software for these types of variants will require additional benchmarking resources to be developed. These may come from additional well-studied reference materials, studying very large cohorts with WGS, aggregating evidence from individual case reports, or benchmarking using cancer genome data. It will be an important goal to increase the number of validated copy number neutral structural variants in germline DNA to improve benchmarking of this class of genetic variant.

*ClinSV* was able to identify all pathogenic variants that were detected using MA by the two clinical laboratories participating in this study. The increased resolution of WGS compared to MA enables *ClinSV* to resolve large CNV with much higher precision, as well as identify thousands of CNV smaller than the resolution of MA, in addition to copy number-neutral SV. Absolute DOC measurements from WGS provide an advantage over relative quantification used in aCGH to accurately resolve the copy number level of regions frequently altered in the general population, often co-coinciding with multi-allelic CNV, which can include genes that are relevant to disease [58]. In support of this high clinical utility, *ClinSV* has been used in a number of research studies, which have identified short, pathogenic CNV affecting single exons [11], overlapping whole-gene deletion-duplications [10], and in adult patients with mitochondrial disease (Davis *et al, in prep*).

By applying *ClinSV* to an unselected clinical cohort within a clinically accredited diagnostic laboratory we have demonstrated the increased clinical utility of CNV calling from WGS data over that of MA. Of particular interest are the 7/16 likely pathogenic and pathogenic CNVs that would not be readily detectable by MA. One could argue that reducing the minimum number of consecutive probes to make a diagnostic call by MA could help resolve some of these CNVs, however this would still only recover at most an additional two CNVs (ID 4, 11) at the cost of significantly increasing the false positive detection rate of the assay. These small CNVs would be intractable to genome-wide MA techniques due to size and due to the heterogeneity of the disease within this cohort beyond the scope of targeted assays such as MLPA. An additional, underappreciated feature of expanding the variant types detectable by WGS is the consolidation of different variant types into a single assay. Cases 3, 5, 13 and 22 show a compound heterozygous state where one affected allele is a CNV and the other is a single nucleotide variant. Depending on the heterogeneity of the presenting disorder, the availability of previous test results and the reporting policies of the laboratory it is plausible that these variants, if detected by different methods at different laboratories may be interpreted as single hits in a recessive disorder, and be overlooked and considered merely a carrier status rather than a diagnostic result when taken as a whole. Most demonstrative of this are the variants identified in *ERCC5* in case 5 where the presenting condition was complex and heterogeneous, the CNV was well below the resolution of MA and (to our knowledge) there are no commercially available MLPA kits to test for such a deletion. Without simultaneous SNV and CNV detection by WGS this patient is likely to have remained undiagnosed. While there have been limited studies showing that WGS performs similarly well compared to MA [20], to our knowledge this is the first publication showing clear clinical utility for genome wide CNV detection at higher resolution than MA.

The ability for *ClinSV* to identify short CNVs from WGS would represent an insurmountable interpretive challenge for clinical laboratories and researchers, without a way to differentiate the potentially pathogenic events from the thousands of polymorphic CNV present in all healthy individuals [12]. A critical advance of *ClinSV* is the use of matched population control data, with five complementary measures of PAF to prioritise rare CNV. The PAF measures derived from *ClinSV* calls in control (PAFV), gnomAD (PAFG), and the 1000-genomes project (PAF1KG) highlight the allele frequency of well-resolved CNV. Perhaps more importantly, the PAF developed from SR and DP raw data highlights genomic regions with ambiguous short-read-mapping, which are depleted from final CNV and SV calls, making this a powerful way to filter variants and common artefacts. Large CNVs affecting multiple genes can be interpreted as they currently are by cytogenetics and research laboratories, whereas smaller candidate CNVs should be assessed in the context of which genes are affected, the patient’s phenotype and family history, known gene-phenotype correlations and sensitivity to gene dosage and haploinsufficiency. While recent recommendations [59,60] will help to standardize this interpretation, it should be noted that the clinical interpretations in the issued pathology reports were performed prior to the publication of these guidelines and were instead based on a combination of previous guidelines [49,50]. When combined with segregation analysis using additional family members, there are typically fewer than 10 variants to inspect.

We have also shown that short-reads are limited in their ability to resolve a subset of SV with repeated sequences surrounding each breakpoint, particularly if no DOC evidence exists (as for copy number-neutral SV) or if the DOC is not sufficient on its own (as for CNV <10 kb). The most frequent repeat class found at breakpoints were tandem repeats and we hypothesize that these repeats may act as a catalyst to form SVs similarly as has been suggested for segmental duplications [61]. Long-read sequencing technologies including Oxford Nanopore and PacBio offer improved performance over repetitive elements, and aberrant GC%, and they can identify thousands of SV missed by short-read sequencing [43]. Long-read sequencing also enables complementary ways of detecting SV by using *de novo* assembly approaches, compared to the reference genome [62]. Furthermore, optical mapping is able to identify even larger SV often missed by current long-read sequencing technology [63]. Today however, for most clinical and research laboratories seeking a single test that can accurately identify all SNVs, indels, and most CNVs and SVs from a patient’s genome, long-read sequencing remains cost prohibitive at the sequencing depths required to also obtain robust SNV and indel performance. Hybrid approaches, combining high-quality short-read sequencing, supplemented with limited amounts of long-read sequencing for CNV detection may become a promising approach in the clinic.

## Conclusion

*ClinSV* can accurately identify pathogenic CNVs and copy number-neutral SVs from short-read WGS data in the size range from 50 bp up to whole-chromosome aneuploidy, at a resolution far higher than existing clinical microarrays. Through its integration, filtration, annotation and visualization framework, it accurately identifies high-quality CNVs and SVs from patient clinical genome data and presents the data in a clinician-and researcher-friendly output. We have demonstrated that *ClinSV* combined with 30– 40x short-read WGS provides increased clinical utility over microarray, resolving an additional 4.7% of cases over the usual standard combination of WES/WGS and microarray. By combining WGS with clinical-grade methods for SNV, indel, CNV and SV detection, it represents a comprehensive, single-test capable of diagnosing a larger portion of genetic disorders than even a combination of previous best methods.

## List of Abbreviations

aCGH: comparative genomic hybridization array
CGH: comparative genomic hybridization
CNV: copy number variant(s)/variation(s)
DOC: depth of coverage
DP: discordant pair(s)
GIAB: Genome in a bottle
indel: short insertion or deletion variant/variation
MA: microarray
MQ: mapping quality
NATA: National Association of Testing Authorities, Australia
PAF: population allele frequency
QC: quality control
SNP: single nucleotide polymorphism
SNV: single nucleotide variant(s)/variation(s)
SR: split read(s)
SV: structural variant(s)/variation(s)
WGS: whole genome sequencing

Other abbreviations see Suppl. Table 2

## Declarations

### Ethics approval and consent to participate

Patient’s MA data and full variant call set was consented for validation experiments in this study. We do not have appropriate patient consent to deposit patient WGS data into public databases.

## Consent for publication

Not applicable

## Availability of data and materials

The *ClinSV* software, installation and usage instructions, its supporting resource files, as well as Singularity and Docker images are available on GitHub (https://github.com/KCCG/ClinSV). The installation requirements are listed on the GitHub link above. The software is implemented in Perl and has been tested on Linux CentOS 6.10 and Ubuntu 14.04. The software is released under an open source license for academic use, and for commercial or diagnostic use, please contact the corresponding authors.

## Authors’ contributions

AEM designed and developed the method including the bioinformatics pipeline. BL called microarray variants and contributed to the design of validation experiments. LB authorized the clinical diagnostic pathology reports, and contributed to the design of the validation experiments. TO and BL performed the MLPA validation. BL, LB and NS contributed with the clinical case study. MJC and MD conceived the project, established and maintained core infrastructure. MJC, MD and SK oversaw the project and contributed ideas. AEM, MJC and BL wrote the manuscript with contributions from LB and SK. AZ contributed patient samples and matched microarray data. GBP provided microarray data and curated pathogenicity calls. GBP, BL and TR provided clinical cytogenetics expertise. MP and DMT provided genomic data from MGRB controls. All authors read and approved the manuscript.

## Acknowledgments

We thank the Kinghorn Centre for Clinical Genomics for assistance with production and processing of genome sequencing data. This research was undertaken with the assistance of resources and services from the National Computational Infrastructure, which is supported by the Australian Government. We thank Australian Clinical Labs and Genome.One for providing the summary of clinically reported CNVs. Thank you to Alexander Drew for manuscript review. We thank Ryan Taft and Michael Eberle from Illumina, and Mike Field from Genetics of Learning Disability (GOLD) for helpful discussions. We thank Pauline Dalzell and Michael F Buckley at the NSW Health Pathology (Prince of Wales SEALS campus) Genetics Laboratory for providing MA data and matched patient DNA. We thank the ASPREE and 45-and-up study coordinators for patient recruitment for the MGRB.

## Supplemental information

Suppl. Data 1 (docx): Supplemental Tables and Figures

Suppl. Data 2 (xlsx): Sample output rare gene affecting variants NA12878 Suppl. Data 3 (pdf): Sample automated QC report NA12878

